# OpenChart-SE: A corpus of artificial Swedish electronic health records for imagined emergency care patients written by physicians in a crowd-sourcing project

**DOI:** 10.1101/2023.01.03.23284160

**Authors:** J Berg, CO Aasa, B Appelgren Thorell, S Aits

## Abstract

Electronic health records (EHRs) are a rich source of information for medical research and public health monitoring. Information systems based on EHR data could also assist in patient care and hospital management. However, much of the data in EHRs is in the form of unstructured text, which is difficult to process for analysis. Natural language processing (NLP), a form of artificial intelligence, has the potential to enable automatic extraction of information from EHRs and several NLP tools adapted to the style of clinical writing have been developed for English and other major languages. In contrast, the development of NLP tools for less widely spoken languages such as Swedish has lagged behind. A major bottleneck in the development of NLP tools is the restricted access to EHRs due to legitimate patient privacy concerns. To overcome this issue we have generated a citizen science platform for collecting artificial Swedish EHRs with the help of Swedish physicians and medical students. These artificial EHRs describe imagined but plausible emergency care patients in a style that closely resembles EHRs used in emergency departments in Sweden. In the pilot phase, we collected a first batch of 50 artificial EHRs, which has passed review by an experienced Swedish emergency care physician. We make this dataset publicly available as OpenChart-SE corpus (version 1) under an open-source license for the NLP research community. The project is now open for general participation and Swedish physicians and medical students are invited to submit EHRs on the project website (https://github.com/Aitslab/openchart-se). Additional batches of quality-controlled EHRs will be released periodically.

## Introduction

Electronic health records (EHRs) are increasingly used around the world to collect background information, clinical observations and measurements and treatment details for each a patient. They are therefore a rich source of information for medical research and public health monitoring and can for example be used to identify diagnostic markers, risk factors or outcome predictors. EHRs could also be connected to medical information systems that support health care professionals and hospital managing staff in their work [1].

To enable the use of EHRs at large scale, automated information extraction is necessary. This is relatively straight-forward for structured EHR data such as laboratory results. However, a substantial part of EHR data consists of unstructured text written by various health care professionals. Natural language processing (NLP), a form of artificial intelligence that has made enormous progress in the last decade, can help to extract information from this unstructured text [1-5]. To develop and validate NLP tools, collections of texts (so-called corpora) derived from EHRs are required. However, due to the sensitive nature of EHR data, it is difficult to access and cannot be shared freely unless sensitive information has been removed, which is time-consuming and costly. Consequently, only a few corpora derived from EHRs have been made publicly available and the vast majority of these is in English, with a few datasets available for other major languages such as French, Spanish and Chinese [1,3,5,6]. The scarcity of publicly available EHR-derived text, especially in less commonly spoken languages, is one of the greatest hinderances for the development of clinical NLP tools and even though significant progress has been made in the English language many other languages still lag behind [2,3,5,7].

For Swedish, some progress in clinical NLP research has been made using corpora with restricted access [8,9]. However, there is, to our knowledge, currently no publicly available corpus with Swedish EHR-derived texts. Even when no publication of the data is intended, getting access to EHR data can take years after ethical approval, even for researchers at Swedish universities. This restricted availability of EHR-derived text data remains a major obstacle for clinical NLP research in Sweden. Fortunately, artificial data can be used as substitute in NLP research to a large extent, as long as the writing style is highly similar to real texts. We therefore set up the “OpenChart-SE” citizen science project in which Swedish physicians and medical students submit artificial EHRs for imagined patients whom they could plausibly encounter at a Swedish emergency department. These artificial EHRs are written in the same language style and structure that is normally used in Swedish hospitals. With this article, we publish the first batch of 50 artificial EHRs from the pilot phase of the OpenChart-SE project, which passed review by experienced physicians, as an open-source resource for Swedish clinical NLP research [10]. Both academic and non-academic NLP researchers can access this resource freely. The OpenChart-SE project has now proceeded to the main data collection phase and we invite Swedish physicians and medical students to submit EHRs through the form on the project website (https://github.com/Aitslab/openchart-se). We will continue to release incoming data after quality control.

## Material and methods

### Data collection

Swedish physicians and medical students, who have experience in writing EHRs, were recruited as citizen scientists for the OpenChart-SE project and asked to write artificial EHRs for imagined but plausible patients at a Swedish emergency department. For data entry, a Swedish mock EHR form was created with KoboToolbox (https://www.kobotoolbox.org/) and published on a public web site (https://github.com/Aitslab/openchart-se) together with submission instructions and a project description. The form was written in Swedish, and fields were accompanied by brief explanations and examples of suitable text (also in Swedish). The fields were based on those used in EHRs in emergency departments in the hospitals of the region of Scania in Southern Sweden, which closely resemble EHRs used in the rest of Sweden. The displayed exam field changed, depending on whether the “priority one patient” option, i.e. patient in need of immediate medical attention, was selected or not. The following fields were present on the form (translation provided here, original Swedish form available on Zenodo [10]):

> *Chief complaint* Primary reason for visiting according to the Rapid Emergency Triage and Treatment System (RETTS)
>
> *Background:* Gender, Age, Social situation, Prior medical conditions, Current circumstances prompting the emergency care visit (“Aktuellt”), Hereditary information, Smoking, Priority one patient
>
> *Exam* (only visible when the priority patient one option was ***not*** chosen): General appearance, Heart, Lung, Abdomen, Systolic blood pressure, Diastolic blood pressure, Saturation, Pulse, Temperature, Breathing frequency, Local status, Ears, Mouth and throat, Neurology
>
> *Exam priority one patient* (only visible when the priority one patient option was chosen): A (Airway), B (Breathing), C (Circulation), D (Disability), E (Exposure)
>
> *ECG and laboratory results*: Laboratory results, ECG results
>
> *Assessment and diagnosis*: Assessment summary, Plan, Main diagnosis (ICD-10 code)

In addition, the form contained an initial section in which participating citizen scientists could submit their contact details and level of professional experience.

In the pilot phase of the project (November 2021 – December 2022), 50 artificial EHRs were collected to assess project feasibility. All 50 artificial EHRs passed quality control and are published as OpenChart-SE corpus, version 1 (Supplemental data 1) in the Zenodo repository [10].

The project is opened for general submission from January 2023.

## Data quality control

All collected artificial EHRs were reviewed by a resident physician from the emergency department at Skåne University Hospital Malmö, Sweden. To pass quality control, both writing style and content had to be realistic, i.e. matching the structure and linguistic characteristics of Swedish emergency department EHRs and describing a plausible Swedish emergency department patient.

## Data exploration

Data exploration was performed in a notebook run Jupyter lab (version 3.2.5) (Supplemental data 2) using Python (version 3.9.5) and its pandas (version 1.3.4) and matplotlib (version 3.5.2) libraries.

Main diagnoses were reviewed manually and synonymous entries harmonized before counting their occurrences.

## Results and discussion

### Design of the OpenChart-SE citizen science platform for collecting artificial Swedish emergency care EHRs from physicians and medical students

To address the severe difficulties in accessing Swedish EHRs for NLP research, we set up the OpenChart-SE project, in which volunteer Swedish medical students and physicians generate artificial EHRs that can be released publicly after quality control (Figure 1).

**Figure 1.**
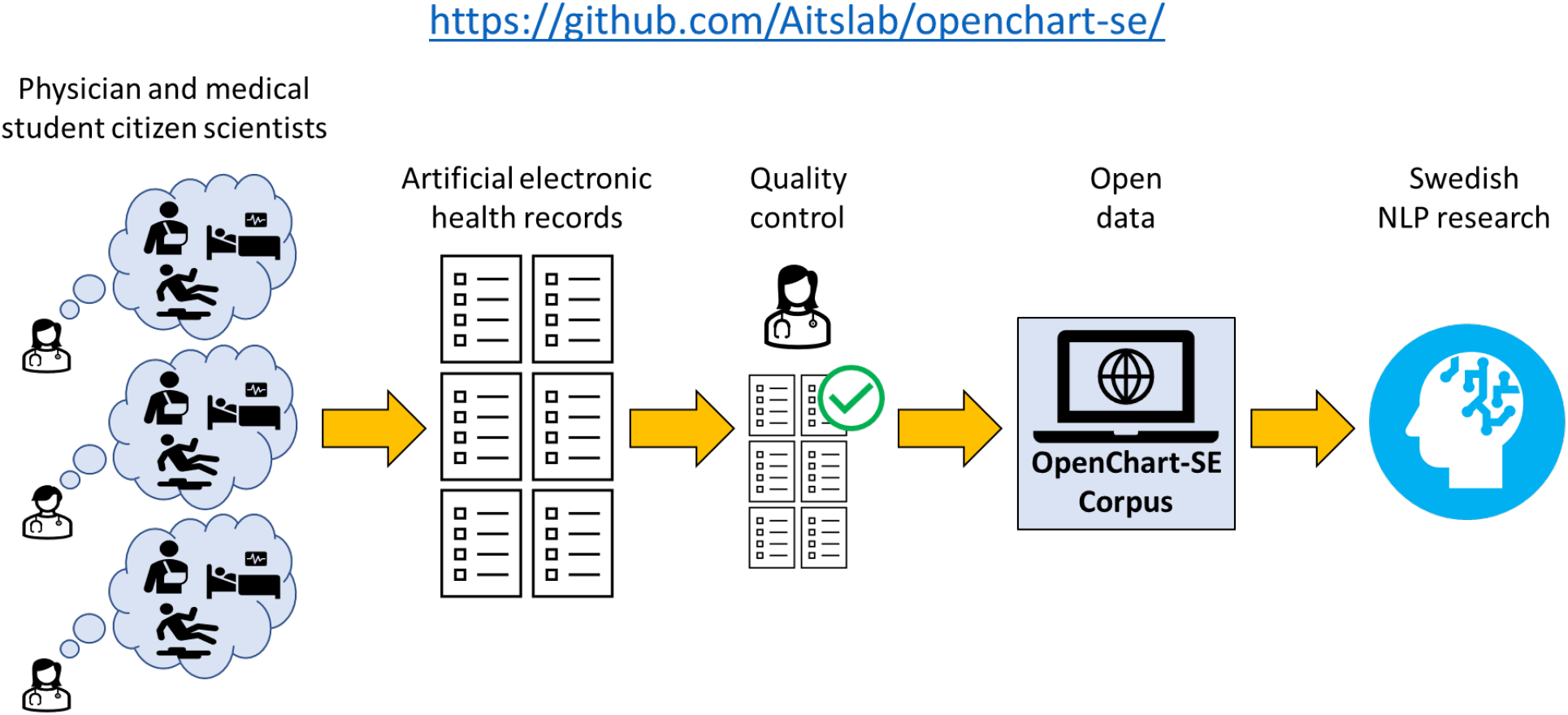
Overview over the OpenChart-SE citizen science project

For data collection, we created a mock EHR form [10] written in Swedish that mimicked an EHR software used in emergency hospital care in the region of Scania, Sweden. Other Swedish regions use highly similar EHRs. We chose to imitate EHRs from emergency care as this area of medicine has a very broad variety of patients and all physicians rotate through it in their training, making them familiar with the style of emergency care EHRs.

The first section of the form was optional and allowed the user to choose between anonymous and non-anonymous data submission. For non-anonymous data submission, Name, Email address and consent for storing and processing this information for the purpose of the project was collected.

The next section required the user to declare their level of medical experience with the options being medical student (semester1-5), medical student (semester 6-11), physician before legitimation, physician after legitimation, resident physician/specialist in emergency care, resident physician/specialist in other medical area.

After this, the fields corresponded to those found in a Swedish emergency care EHR system: *Chief complaint (“Sökorsak”), Background (“Bakgrund”), Exam (“Status/Status larm”), ECG and laboratory results (“EKG och lab” and Assessment and diagnosis (“Bedömning och åtgärd”)*. One of two alternative fields (“Status larm” or “Status”) was displayed for the exam section, depending on whether the patient was classified as priority one patient (“larmpatient”) in the background section or not. Priority one patients are patients in need of immediate medical attention who receive a structured clinical examination according to the ABCDE scheme (Airway, Breathing, Circulation, Disability, Exposure) with the goal of identifying threats to life as quickly as possible. The “Status larm” field follows this scheme.

The form was published on https://github.com/Aitslab/openchart-se. Project feasibility and clarity of the instructions was evaluated during a 1-year pilot phase. During this pilot phase, we collected 50 artificial EHRs, primarily written by physicians and medical students who had been recruited as citizen scientists through personal contacts. The form was also open for general participation but for the pilot phase no public announcement was made. All 50 artificial EHRs were deemed realistic by an experienced emergency care physician. They are released as OpenChart-SE corpus (version 1) in the Zenodo repository [10]. Due to the success of the pilot phase, the project has now proceeded to its main phase, and we welcome all physicians and medical students who are trained in writing Swedish EHRs to submit additional artificial EHRs on https://github.com/Aitslab/openchart-se. After quality control, this additional data will be released periodically to expand the OpenChart-SE corpus.

### The OpenChart-SE corpus covers a broad variety of emergency care scenarios

The first batch of 50 artificial EHRs (OpenChart-SE corpus, version 1) were explored in detail. All were plausible in writing style and patient characteristics, resembling real Swedish emergency department EHRs. Text length varied between 954 and 2738 characters (Figure 2).

**Figure 2.**
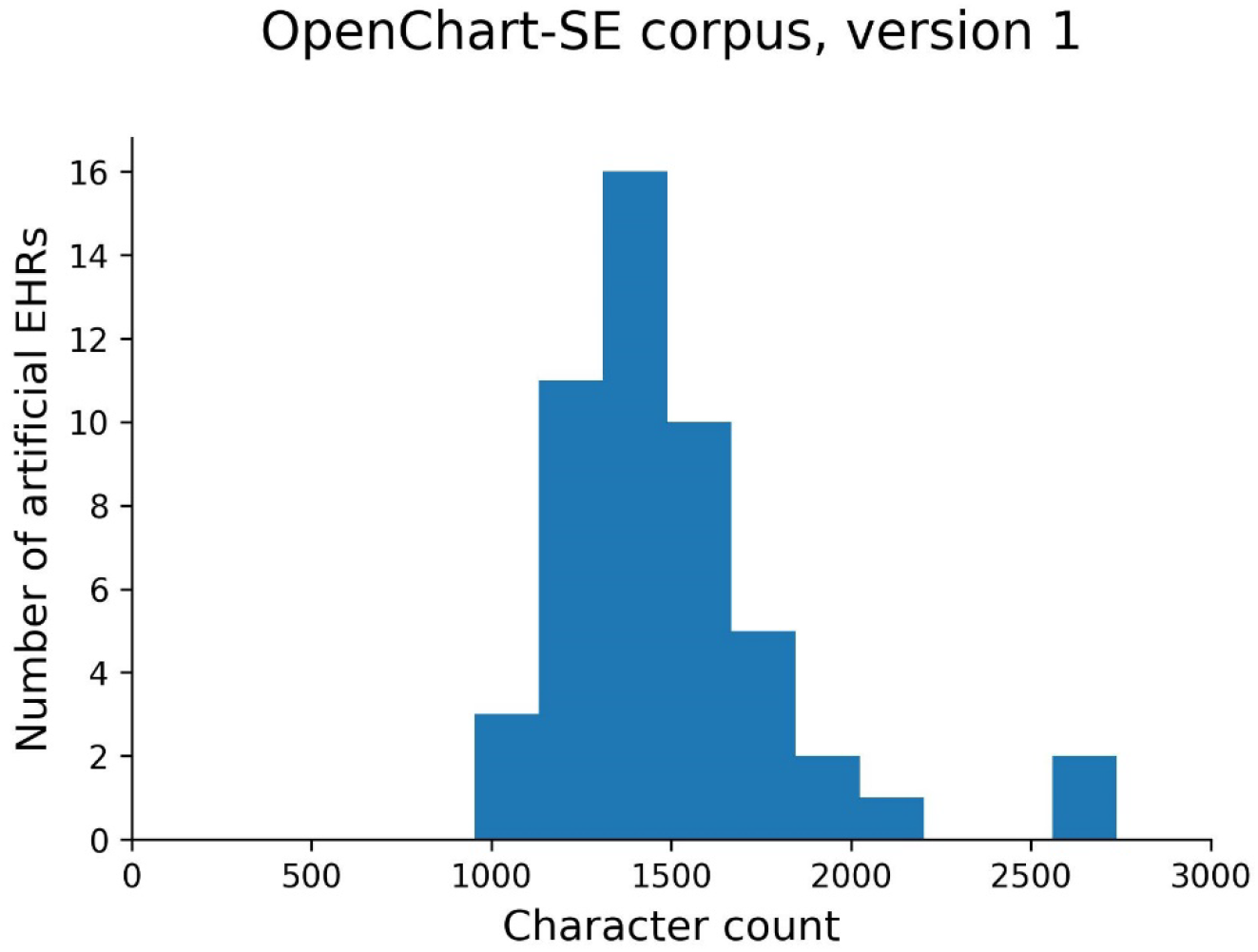
Length of artificial EHRs in the OpenChart-SE corpus, version 1.

Six artificial EHRs (12%) described imagined priority one patients. 29 artificial EHRs (58%) described imagined female patients and 21 imagined male patients, whereas there were no artificial EHRs in which the option “other sex” had been chosen. The age of the imagined patients varied from 18 to 94 years, with a mean of 52 years (Figure 3). The lack of pediatric patients in the dataset reflects reality as children are typically handled in separate pediatric emergency departments in Sweden.

**Figure 3.**
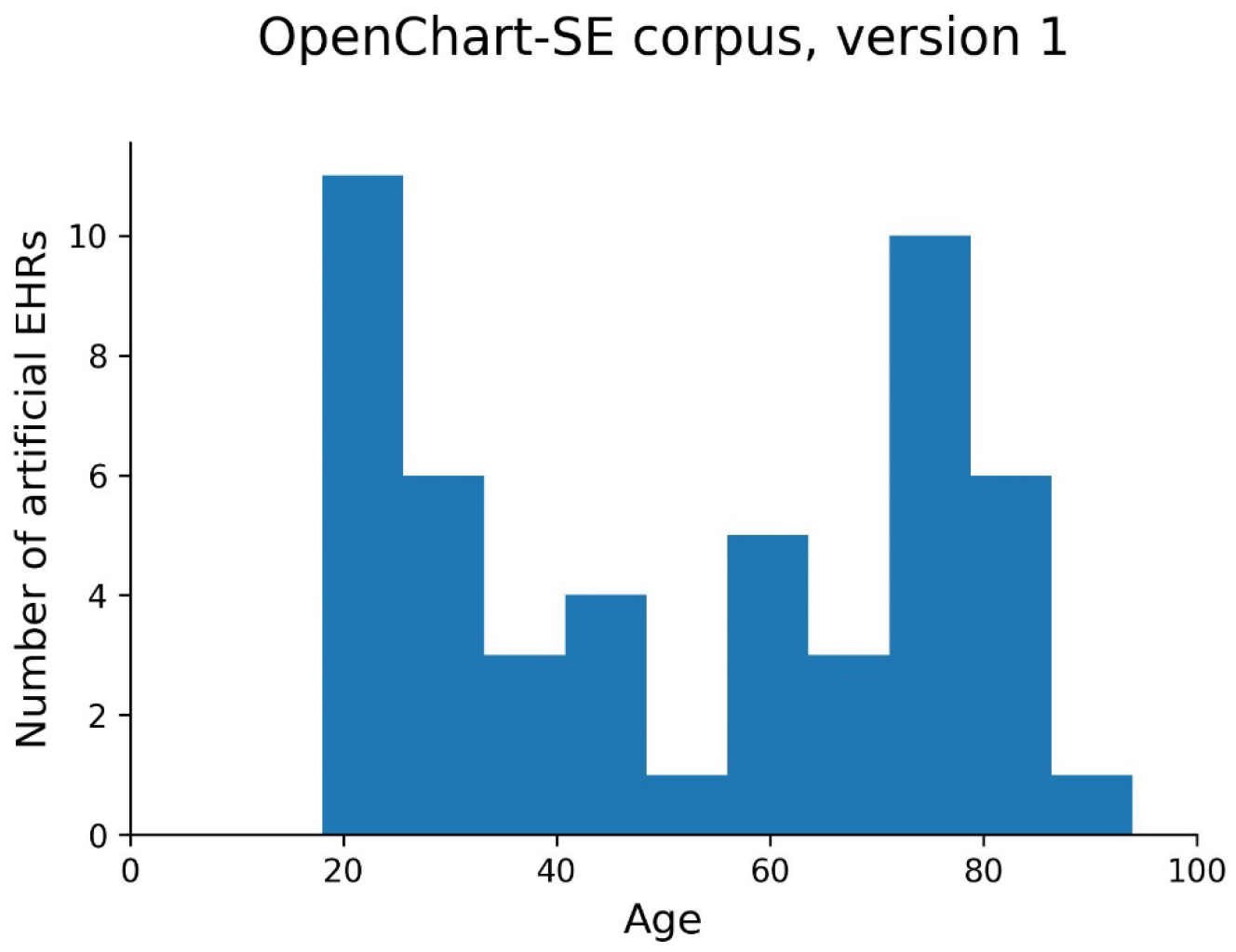
Age distribution of imagined patients in the OpenChart-SE corpus, version 1.

As is typically seen in real EHRs, all artificial EHRs lacked text in at least one field (indicated as “NULL”), including one artificial EHR with a missing main diagnosis. For further analysis, all main diagnoses were reviewed, and synonymous entries harmonized. In the files of the published corpus the text remains unchanged, however, to reflect real linguistic variation and errors. 32 artificial EHRs had a unique main diagnosis and 17 a main diagnosis described in multiple artificial EHRs. The most frequent main diagnoses were “Other and unspecified abdominal pain” (ICD-10 code R10.4; in 4 EHRs), “Calculus of ureter” (ICD-10 code N20.1; in 3 EHRs) and “Dyspnea” (ICD-10 code R06.0, in 3 EHRs).

## Limitations

This corpus does not contain real EHRs as they are too sensitive to be released publicly and therefore no clinical insights can be derived from it. However, the artificial EHRs closely resemble real Swedish emergency department EHRs in linguistic style and medical content as they were written by Swedish physicians and medical students and validated by an experienced emergency care physician. The dataset is thus highly suited as training and validation data for clinical NLP tools.

## Conclusions

In conclusion, the first version of the OpenChart-SE corpus contains 50 artificial Swedish EHRs describing a broad variety of imagined adult emergency care patients. As the artificial EHRs were written and validated by Swedish medical professionals they closely resembled real Swedish EHRs. As real Swedish EHRs are extremely difficult to access, this dataset can serve as a key resource for Swedish clinical NLP research.

All physicians and medical students with training in writing EHRs are welcome to submit artificial EHRs through https://github.com/Aitslab/openchart-se. Updated versions of the corpus with additional quality-controlled artificial EHRs will be released periodically.

## Data Availability

All data produced are contained in the manuscript or available online athttps://doi.org/ https://zenodo.org/record/7499831.

https://doi.org/ https://zenodo.org/record/7499831

https://github.com/Aitslab/openchart-se

## Acknowledgement

We thank all citizen scientists for contributing artificial EHRs and all members of our research groups, who provided helpful comments throughout the development of this project.

This study was supported by a grant to Science for Life Laboratory from the Knut and Alice Wallenberg (KAW) Foundation (S.A. 2020.0182), which was distributed through the SciLifeLab and KAW National COVID-19 Research Program. The project is conducted in the AI Lund research environment at Lund University.

## Competing interests

The authors have no competing interests related to this publication.

## CRediT author statement

Johanna Berg: Conceptualization, Methodology, Software, Validation, Investigation, Resources, Data Curation, Writing – Review & Editing, Supervision, Project administration

Carl Ollvik Aasa: Methodology, Software, Writing – Review & Editing

Björn Appelgren Thorell: Data Curation, Validation, Writing – Review & Editing

Sonja Aits: Conceptualization, Methodology, Software, Validation, Formal analysis, Investigation, Resources, Data Curation, Writing - Original draft, Writing – Review & Editing, Visualization, Supervision, Project administration, Funding acquisition

## Supplemental data

Supplemental data 1. OpenChart-SE corpus, version 1. Collection of 50 artificial EHRs (unprocessed text). Available from Zenodo [10] (released 2022-12-19).

Supplemental data 2. Jupyter notebook with code for data exploration

## Notes

### Competing Interest Statement

The authors have declared no competing interest.

