## Supplemental data 2 for "OpenChart-SE: A corpus of artificial Swedish electronic health records for imagined emergency care patients written by physicians in a crowd-sourcing project": suppl_data_3_openchart-se_dataexploration.ipynb

```
# %% [markdown]
# Corpus version 1, data exploration

# %% [markdown]
Notebook author: Sonja Aits, sonja.aits [at] med.lu.se

# %% [code]
import os
import re
import pandas as pd
import matplotlib.pyplot as plt

# %% [code]
# Data exploration and extraction for the set of EHR .txt files present in the "inputfolder"

def analysis(inputfolder):
    # get list of filenames in folder
    filenames = os.listdir(folder)
    
    # create counter for priority one patients
    larm_count = 0
    
    # write headings to csv output file            
    outputfile = folder + 'analysis.csv'
    with open(outputfile, 'a') as f:
        f.write('filename'+';'+'number_of_characters'+';'+'priority status'+';'+'diagnosis'+';'+'sex'+';'+'age'+'\n')
    
    #iterate over EHR files to extract statistics    
    for file in filenames:
        if file != 'Readme.md':
            
            # open file and read file content
            path = folder + file
            doc = open(path, "r", encoding='utf-8') 
            data = doc.read()
            
            
            # count characters in file
            number_of_characters = len(data)-47 #get the character count; 47 characters from the validation status line subtracted
            
            
            # extract gender information
            pattern = '(?<=kon:\s)[A-Za-z]+'
            sex = re.findall(pattern, data)
            sex = str(sex)
            sex = sex.replace('[', '')
            sex = sex.replace(']', '')
            sex = sex.replace('\n', '')
            sex = sex.replace('\'', '')
            
            
            # extract age
            pattern = '(?<=alder:\s)[\d]+'
            age = re.findall(pattern, data)
            age = str(age)
            age = age.replace('[', '')
            age = age.replace(']', '')
            age = age.replace('\n', '')
            age = age.replace('\'', '')
            
            
            # extract "larm" status                    
            if 'larm: ja' in data:
                larm = 'larm: ja'
                larm_count = larm_count + 1
            
            else:
                larm = 'larm: nej'
                
         
            # extract main diagnosis
            pattern = '(?<=huvuddiagnos_001:\s)[\d\wåäöÅÄÖ .,]+'
            diagnosis = re.findall(pattern, data)
            diagnosis = str(diagnosis)
            diagnosis = diagnosis.replace('[', '')
            diagnosis = diagnosis.replace(']', '')
            diagnosis = diagnosis.replace('\n', '')
            diagnosis = diagnosis.replace('\'', '')
            
            
            doc.close()
            
            
            # write extracted information to csv output file            
            outputfile = folder + 'analysis.csv'
            with open(outputfile, 'a') as f:
                f.write(file + ';' + str(number_of_characters) + ';' + larm + ';' + diagnosis + ';' + sex + ';' + age +'\n')
            
                
    print("EHRs with priority one patients: "+ str(larm_count))

# %% [code]
folder = INPUTPATH # ADD PATH TO YOUR INPUT FOLDER HERE
analysis(folder)

# %% [code]
# Load extracted information into dataframe for further exploration
csvfile = CSVPATH # ADD PATH TO YOUR ANALYSIS.CSV FILE HERE
df = pd.read_csv(csvfile, encoding='latin', sep = ';')
df.head(50)

# %% [code]
# get statistics for number of characters
# if max count >3000 the upper limit for the x-axis in the character count histogram needs to be adjusted
df['number_of_characters'].describe()

# %% [code]
# get statistics for sex
df['sex'].describe()

# %% [code]
# get statistics for age
# if max age >100 the upper limit for the x-axis in the age histogram needs to be adjusted
df['age'].describe()

# %% [code]
# Visualize distribution of variable "age" in dataset

# Create a figure instance
fig = plt.figure()

# Create an axes instance
ax = fig.add_subplot(111)

# Remove right and top lines around plot
right = ax.spines["right"]
right.set_visible(False)
top = ax.spines["top"]
top.set_visible(False)

# Create the histogram
ax.hist(df['age'])

# Add axis labels and header
plt.xlabel('Age', fontdict = {'fontsize': 13})
plt.ylabel('Number of artificial EHRs', fontdict = {'fontsize': 13})
plt.title('OpenChart-SE corpus, version 1', fontdict = {'fontsize': 16}, y=1.1)

# Define x-axis range
plt.xlim([0, 100]) #upper limit needs to be adjusted if patients over 100 years were in the dataset

# Save and show plot
jpg1 = OUTPUT PATH # ADD DESIRED OUTPUT PATH FOR JPG FILE 
plt.savefig(jpg1, dpi = 300, bbox_inches='tight')
plt.show()
plt.close()

# %% [code]
# Visualize distribution of character count in dataset

# Create a figure instance
fig = plt.figure()

# Create an axes instance
ax = fig.add_subplot(111)

# remove right and top lines around plot
right = ax.spines["right"]
right.set_visible(False)
top = ax.spines["top"]
top.set_visible(False)

# Create the histogram
ax.hist(df['number_of_characters'])

# Add axis labels and header
plt.xlabel('Character count', fontdict = {'fontsize': 13})
plt.ylabel('Number of artificial EHRs', fontdict = {'fontsize': 13})
plt.title('OpenChart-SE corpus, version 1', fontdict = {'fontsize': 16}, y=1.1)

# Define x-axis range
plt.xlim([0, 3000])  #upper limit needs to be adjusted if character counts >3000 were in the dataset

#Save and show plot
jpg2 = OUTPUT PATH # ADD DESIRED OUTPUT PATH FOR JPG FILE 
plt.savefig(jpg2, dpi = 300, bbox_inches='tight')
plt.show()
plt.close()

# %% [code]
```
